# Socioeconomic Factors Influencing Breastfeeding Duration in Bangladesh: *An Analysis of the Bangladesh Demographic and Health Survey*

**DOI:** 10.1101/2023.10.15.23297049

**Authors:** Bashirul Alam, Mohammad Abu Tareq Rony, Md Aminul Islam, Md Khalid Hasan, Kanamik Kani Khan

## Abstract

**Background:** This cross-sectional survey aimed to investigate the socioeconomic factors influencing the Duration of Breastfeeding in Bangladesh, using data from the Bangladesh Demographic and Health Survey (BDHS) 2017-18, with a sample of 4,881 mothers having children under two years old. The mean Duration of exclusive Breastfeeding was 15.23 months, with a standard deviation of 9.83 months.

**Methods:** The study utilized statistical tests such as independent sample t-test and one-way analysis of variance (ANOVA) to identify significant differences in the Duration of Breastfeeding between different groups. Pearson’s correlation coefficient was used to explore the linear relationship between the Duration of Breastfeeding and other continuous variables. At the same time, multinomial logistic regression analysis was employed to determine the predictors of breastfeeding duration.

**Result:** The study revealed significant variations in the Duration of exclusive Breastfeeding based on various demographic, health-related, socioeconomic, and anthropometric factors. Women who received antenatal care had a significantly shorter duration of exclusive breastfeeding compared to those who did not receive such care. However, no significant differences were observed in breastfeeding duration between male and female children or between those born via cesarean section versus vaginal delivery. Also, mothers who gave birth in a hospital or clinic breastfed exclusively for a shorter period than those who gave birth at home. Moreover, mothers who worked outside the house had a longer duration of exclusive Breastfeeding than homemakers.

The study also noted substantial differences in exclusive breastfeeding duration based on religion and mothers’ body mass index (BMI). Women in the Barisal region breastfed exclusively for a shorter time than those in other regions. Additionally, women classified as underweight (BMI <18.5 kg/m2) breastfed exclusively longer than those with average weight or obese mothers.

**Conclusion:** Policymakers and program implementers can use these findings to work with specific groups of mothers, mainly those less educated, residing regions, or possessing distinct health or socioeconomic characteristics, to promote optimal infant feeding practices and improve overall child health outcomes by breastfeeding practices in Bangladesh.

**Ethics and consent:** This study does not contain personal identifiable information (PII), so ethical approval is not required. Data collection has been done by obtaining consent.

## Introduction

Breast milk is the best food for newborns because it is specially formulated to meet all their nutritional demands for the first six months of life [1]. Breast milk is highly encouraged to be consumed by newborns since it includes a lot of antioxidants that protect infants from viruses [2, 3]. Antioxidants such as vitamins C and E, which lessen oxidative damage to various human tissues, are vital antioxidants during Breastfeeding [4]. Breastfed infants are generally protected from allergies, illnesses, and obesity [4]. This simultaneously lowers the chance of developing childhood illnesses such as diabetes, cancer, and ear infections [5]. The nutrients in breast milk are naturally balanced properly, delivered in bio-available forms, and are easy to digest [6]. Scientist has been studying the impact of Breastfeeding on baby health and development for decades [7]. According to Hoefer and Hardy’s 1929 study [8], breastfed newborns outperform artificially fed ones regarding cognitive development in childhood. Children who have been breastfed perform better on mental and cognitive tests than children who have not been breastfed [9]. Additionally, Breastfeeding prevents newborns from experiencing constipation, diarrhea, or stomach discomfort and lowers postnatal death rates [10]. It can support better motor and cognitive development [11] and decrease the rates of sudden infant death syndrome [12]. Reduced incidence of type 2 diabetes, ovarian and breast cancers, lactation amenorrhea, which may function as a natural birth control, and appropriate weight recovery are among the maternal advantages of Breastfeeding [13]. Breastfeeding has other benefits besides health advantages, including financial, environmental, and psychological benefits [14]. Additionally, it has been demonstrated that breastfed children have a higher intellectual quotient (IQ)[15].

Breastfeeding is a natural and essential aspect of child-rearing but is often overlooked in many societies. Breast milk is not only a source of nutrition for infants but also provides immunological protection, hormones, enzymes, and growth factors vital for an infant’s healthy development [16]. In Bangladesh, Breastfeeding is a deeply ingrained cultural practice, and it is the preferred method of feeding new-borns [17]. Despite this, many challenges are faced by breastfeeding mothers and infants in Bangladesh, including limited knowledge of proper breastfeeding techniques, cultural stigmas, and lack of support from family and healthcare providers [18]. The breastfeeding period is crucial for both the child and the mother, significantly impacting their health and well-being. Breastfed infants for at least six months have a lower risk of developing respiratory infections, gastrointestinal diseases, and other infectious diseases than formula-fed infants [19]. Breastfeeding also provides long-term benefits, such as lower rates of obesity, diabetes, and hypertension in childhood and adulthood [20].

Breastfeeding has also been shown to have a positive impact on maternal health. Mothers who breastfeed have a reduced risk of postpartum hemorrhage, anaemia, and certain types of cancer, including breast and ovarian cancer [21]. Breastfeeding also promotes bonding through skin-to-skin connection between the mother and the infant and provides emotional benefits for both. However, the Duration of Breastfeeding is critical in determining its health benefits. While exclusive Breastfeeding (Exclusive Breastfeeding - defined as the practice of only giving an infant breast milk for the first six months of life.) for six months is recommended by the World Health Organization (WHO), many mothers in Bangladesh may stop breastfeeding early due to cultural practices, lack of support, and socioeconomic factors [22]. A study conducted in Bangladesh found that only 52% of mothers exclusively breastfed their infants for the first six months [23].

The socioeconomic condition of women in Bangladesh can also affect their ability to breastfeed for an extended period.[1] Women who live in poverty may not have access to proper nutrition or healthcare, which can impact their ability to produce breast milk [24]. Additionally, women who work outside the home may face challenges in managing time and space to breastfeed their infants or express breast milk. To address these challenges, various initiatives have been implemented in Bangladesh to promote and support Breastfeeding. For example, the government has established breastfeeding corners in public places to provide a private and hygienic space for mothers to breastfeed [25]. In addition, healthcare providers have been trained to provide counselling and support to breastfeeding women, and community-based support groups have been formed to provide peer support [17]. The Effects of The Breastfeeding Period on Child Health and Mothers’ Socioeconomic Condition in Bangladesh is a research project that aims to contribute to the existing knowledge on breastfeeding practices and their impact on children and maternal health in Bangladesh. The study will use data from the Bangladesh Demographic and Health Survey (BDHS) 2017-18 to explore the socioeconomic factors influencing the Duration of Breastfeeding in Bangladesh.

The findings of this study will be essential in informing policies and programs aimed at promoting and supporting Breastfeeding in Bangladesh. By identifying the barriers and challenges breastfeeding mothers and infants face, policymakers can design relevant interventions to improve breastfeeding practices and encourage the optimal Duration of Breastfeeding. This, in turn, will help to understand how to obtain improved health outcomes for mothers. The Duration of Breastfeeding plays a crucial role in determining its health benefits, and socioeconomic factors can impact mothers’ ability to breastfeed for an extended period. However, initiatives have been implemented to promote and support Breastfeeding, including establishing breastfeeding corners and training healthcare providers to provide counseling and support to breastfeeding mothers. The study aims to investigate the socioeconomic factors influencing the Duration of Breastfeeding in Bangladesh. The findings will be essential in informing policies and programs to promote and support Breastfeeding.

Breastfeeding tradition is prevalent in Bangladesh in comparison to many other nations. According to UNICEF, 65% of women in Bangladesh exclusively breastfeed their newborns until five months old, the highest in all countries surveyed [26]. As far as we know, a few studies on the Duration of breastfeeding practices in Bangladesh use BDHS-1999-2000 [27] and BDHS-2004 datasets. However, no analysis of the Duration of breastfeeding practices in Bangladesh using the BDHS-2017-18 dataset is available. In this study, we aim to understand how socioeconomic factors of mothers can impact breastfeeding duration and how that links to the health of the new-borns.

## Literature Review

Breastfeeding is the most critical and cost-effective way to improve child health outcomes in developing countries like Bangladesh. It is not only beneficial for the child’s health but also positively impacts the mother’s socioeconomic status. Bangladesh has one of the highest breastfeeding rates in the world [2]. However, still, there is a need to understand the impact of the breastfeeding period on child health and mothers’ socioeconomic conditions. This literature review aims to analyse the existing studies and research articles to understand the effects of breastfeeding duration on child health and mothers’ socioeconomic conditions in Bangladesh.

Breastfeeding is considered one of the most critical factors in reducing infant mortality and morbidity rates. It provides a range of essential nutrients that help build the child’s immune system, making them less prone to diseases. Studies suggest that Breastfeeding decreases the risk of childhood diseases such as diarrhoea, pneumonia, and malnutrition, reducing child mortality rates [20]. The Duration of exclusive Breastfeeding also significantly impacts child health outcomes. A study conducted in rural Bangladesh found that children exclusively breastfed for six months had a lower incidence of acute respiratory infection, diarrhoea, and fever than those not solely breastfed [18]. The study concluded that exclusive Breastfeeding for six months was associated with better child health outcomes.

Breastfeeding has a positive impact on mothers’ socioeconomic conditions in Bangladesh. Studies suggest that Breastfeeding reduces the cost of healthcare for mothers and improves their nutritional status [28]. Breastfeeding also helps mothers return to work faster, reducing the loss of wages and productivity [29]. A study conducted in Bangladesh found that mothers who exclusively breastfed their infants for six months had a lower risk of postpartum depression [30]. Breastfeeding also has a positive impact on the mothers’ household expenditure. A study conducted in rural Bangladesh found that Breastfeeding reduced healthcare costs and increased household disposable income, leading to better socioeconomic conditions [31].

Despite the numerous benefits of Breastfeeding, several challenges hinder the practice of exclusive Breastfeeding in Bangladesh. One of the significant challenges is the lack of knowledge and awareness among mothers about the benefits of exclusive Breastfeeding [18]. Another challenge is the lack of support from family members and healthcare professionals [32]. Furthermore, Bangladesh’s cultural and social norms discourage Breastfeeding in public, making it difficult for mothers to breastfeed outside of their homes [33]. The marketing and promotion of breastmilk substitutes also contribute to declining exclusive breastfeeding practices in Bangladesh [34].

Breastfeeding is critical in improving child health outcomes and positively impacting mothers’ socioeconomic conditions in Bangladesh. The Duration of exclusive Breastfeeding has a significant impact on child health outcomes, and it is associated with a lower risk of childhood diseases. Breastfeeding also reduces healthcare costs for mothers and improves their nutritional status, leading to better socioeconomic conditions. However, several challenges hinder exclusive breastfeeding practices in Bangladesh, including a lack of knowledge and awareness, lack of support from family members and healthcare professionals, cultural and social norms, and marketing of breastmilk substitutes. Addressing these challenges through policy interventions, community-based programs, and education campaigns is essential to promote exclusive breastfeeding practices in Bangladesh.

## Methods

### 3.1 Population and Study Design and

Twenty thousand one hundred twenty-seven married Bangladeshi women between the ages of 15 and 49 participated in the Bangladesh Demographic and Health Survey (BDHS) 2017-18, which gathered data on sociodemographic, health, anthropometric, and lifestyle factors. Data collection started on 24 October 2017 and finished on 15 March 2018. The BDHS-2017-18 collected data on breastfeeding duration (DB) among children born three years before the survey. This study, which included both urban and rural settings, was nationally representative and included all Bangladesh administrative areas (divisions). All the details about the study’s design, target audience, data collection method, tools, data reliability, questionnaire, etc., have already been covered elsewhere [35]. Data from the BDHS-2017-18 were used in the current investigation.

### 3.2 Sampling

The Demographic and Health Surveys (DHS) program is the primary resource for gathering and distributing precise, nationally representative data on population health in developing nations [36]. The survey is based on a two-stage stratified sample of households. In the first stage, 675 EAs were selected with probability proportional to EA size, with 250 EAs in urban areas and 425 in rural areas. In the first stage, the sample was drawn by BBS, following the specifications provided by the DHS team. A complete household listing operation was then carried out in all selected EAs to provide a sampling frame for the second-stage selection of households. In the second sampling stage, a systematic sample of 30 homes on average per EA was selected to provide statistically reliable estimates of key demographic and health variables for the country, for urban and rural areas separately, and for each of the eight divisions. By this design, 20,250 residential households were selected. Completed interviews were expected from about 20,100 ever-married women aged 15–49. In addition, in a subsample of one-fourth of the households (about 7–8 households per EA), all ever-married women age 50 and older, never-married women age 18 and older, and all men age 18 and older were weighed and their height measured. Blood pressure and blood glucose testing were conducted in the same households for all adult men and women aged 18 and older [37].

From the primary dataset, we exclude the respondents who did not answer the question about the breastfeeding period. Besides this, some missing values and incomplete responses were removed from the dataset; eventually, there were 4881 samples for our final dataset.

### 3.3 Study Variables

Our study variables are divided into two parts: Dependent variables and independent variables.

**Table 01:** List of the Variables.

| **Dependent Variable** | **Independent Variables** |  |  |
| --- | --- | --- | --- |
|  | Demographic and related Variables | Socio-economic Variable | Anthropometric variables |
| **Months of Breastfeeding** | Mother's Age | Mother's Education Level | Mother's body mass index |
|  | Antenatal Care visits during pregnancy | Father's education level |  |
|  | Total Child Ever Born | Mother's occupation |  |
|  | Mother's age at 1<sup>st</sup> birth | Geographic position (division) |  |
|  | Sex of the children | Religion |  |
|  | Mode of the delivery | Place of the residence |  |
|  | Place of the delivery | Household wealth quintile |  |

### 3.4 Statistical Analysis

Independent sample t-test and one-way analysis of variance (ANOVA) were used to find the significant difference in Duration of Breastfeeding (DB) between two and more than two groups, respectively. Data were checked for the standard assumptions of independent sample t-tests and ANOVA. The normality and homogeneity of cohort variances were checked using the Kolmogorov– Smirnov nonparametric test, a standard probability plot, and the Levene test, respectively. We used Chi-square to determine whether there is an association between the categorical variables in our data. Pearson’s correlation coefficient was used to find the degree of the linear relationship between the Duration of Breastfeeding and other selected continuous variables. Finally, multinomial logistic regression analysis was used to identify the predictors of the Duration of Breastfeeding.

### 3.5 Instrument

We used R programming language, Stata MP-17 version, and SPSS (Statistical Package for Social Sciences) version 26 to clean the data and perform the analysis, and statistical significance was accepted at p < 0.05.

## Results

**Table 02:**
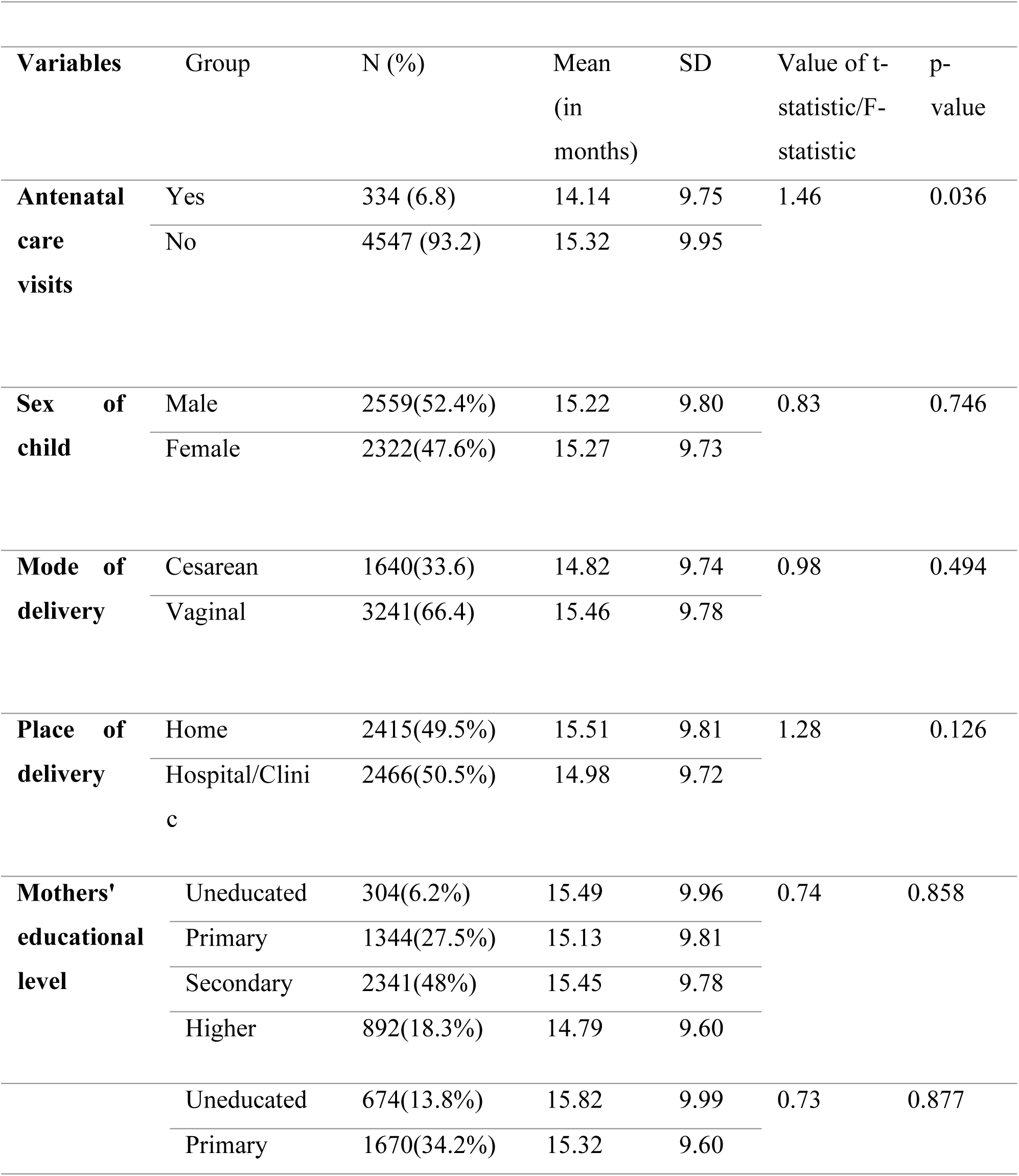
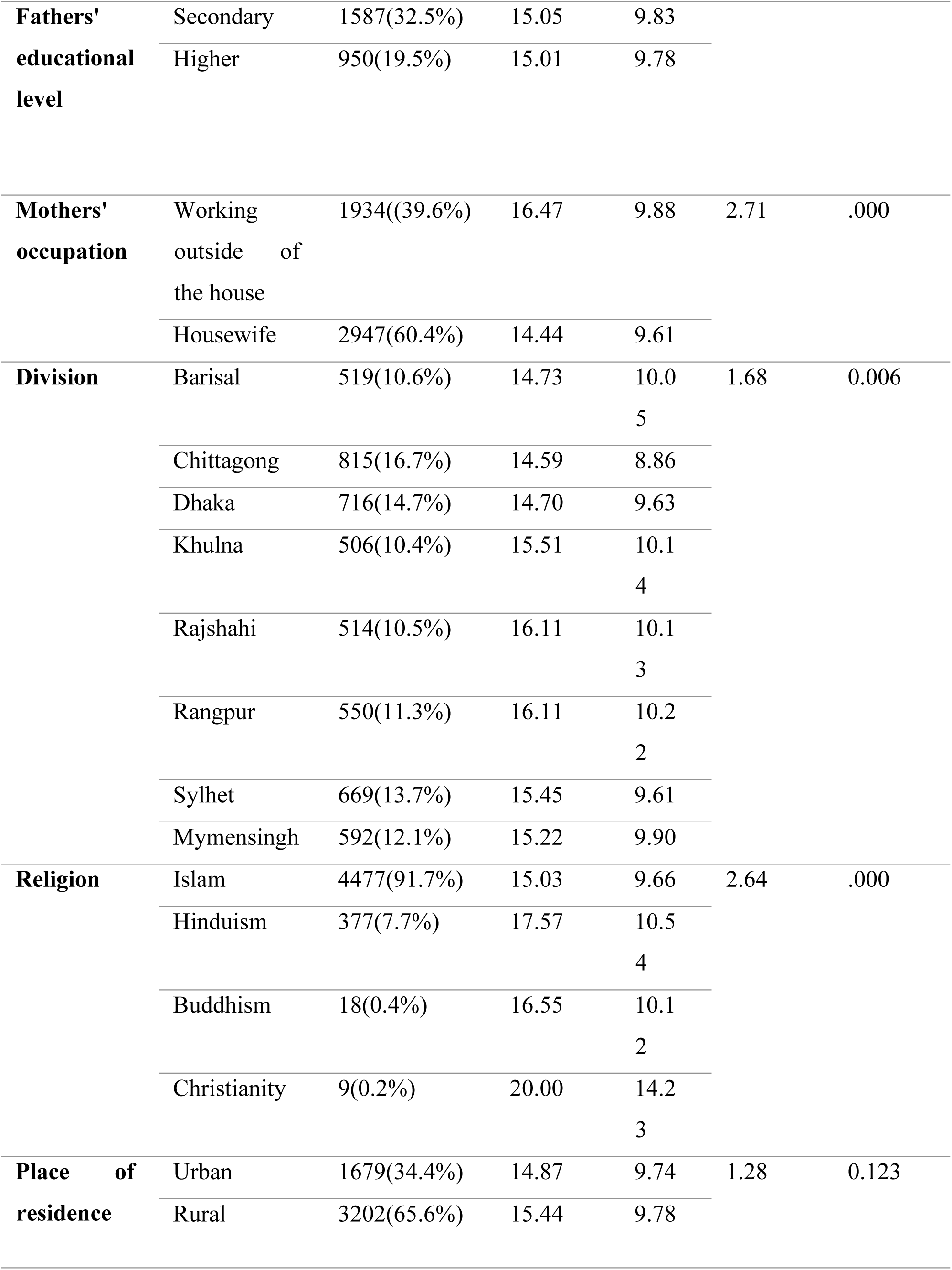
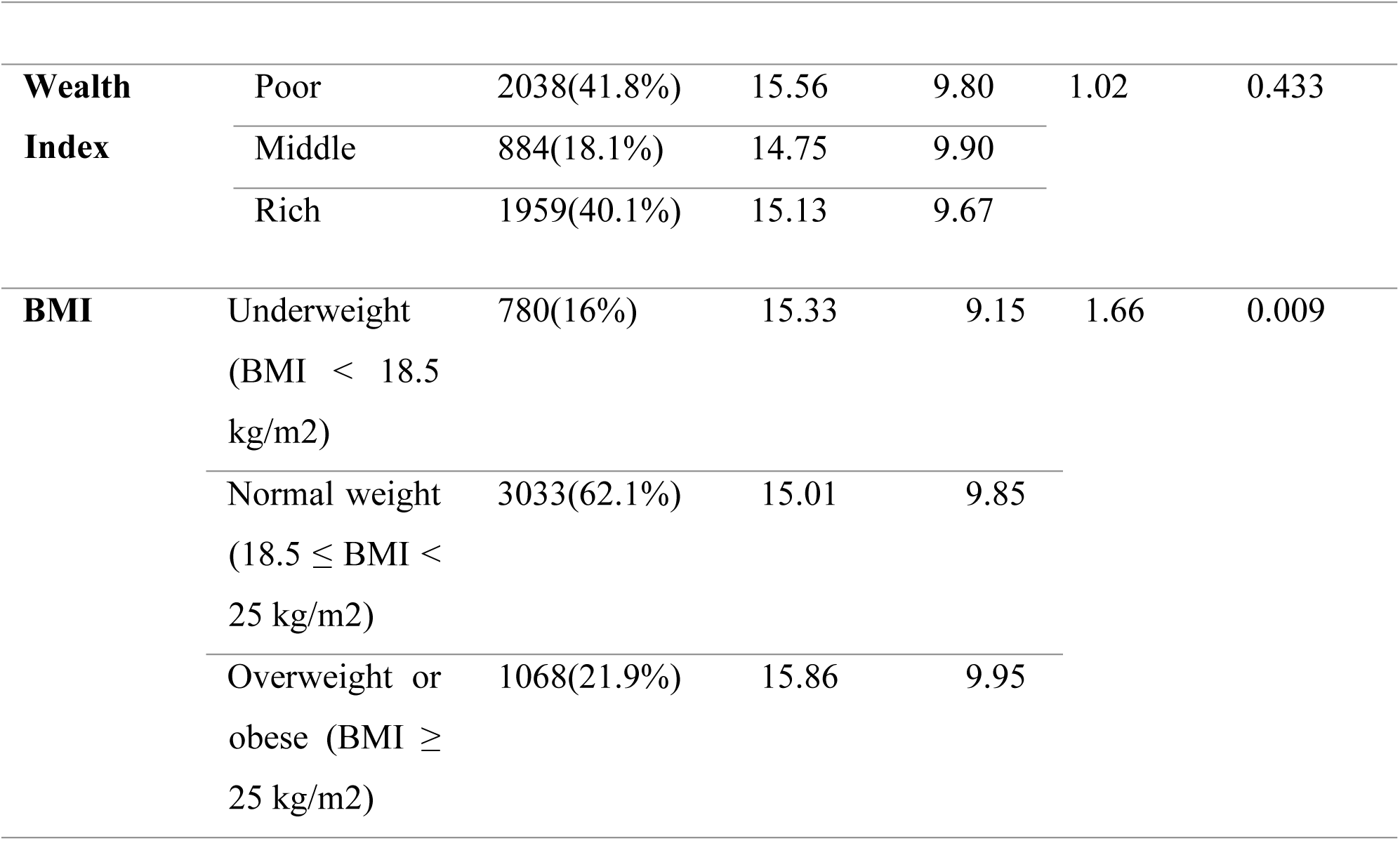
Duration of Breastfeeding among mothers by sociodemographic factors.

The results indicate that mothers who received antenatal care had a significantly shorter duration of exclusive breastfeeding than those who did not. Additionally, there were no significant differences in the Duration of exclusive Breastfeeding between male and female children or between those born via caesarean section versus vaginal delivery.

Women who gave birth in a hospital or clinic had a significantly shorter duration of exclusive breastfeeding than those who gave birth at home. Women who worked outside the house had a significantly longer duration of exclusive Breastfeeding than homemakers.

There were no significant differences in the Duration of Exclusive Breastfeeding by mothers’ or fathers’ educational level, division, place of residence, or wealth index. However, there were significant differences by religion, with women of non-Muslim faiths having a longer duration of exclusive Breastfeeding than Muslim women.

Finally, there were significant differences in the Duration of Exclusive Breastfeeding by mothers’ BMI category, with underweight mothers having the shortest Duration and obese mothers having the most extended Duration.

From the table, we also found that for Antenatal Care visits, the Mother’s occupation, Division, Religion, and Mother’s BMI have a P-value (<.05). We can say that this independent variable has a significant association with the Duration of Breastfeeding.

A chi-square (χ2) statistic is a test used to measure how expectations compare to actual observed data or model results [38]. This test is used to determine whether there is an association between variables.

Let us consider,

H_0_: There is no association between the Duration of Breastfeeding and independent variables (Division, Religion, Mother’s Occupation, Mother’s BMI)

**Table 03:** χ_2_ table for Duration of Breastfeeding.

| Independent Variables | $\chi^2$ value | Degree of Freedom | P-value |
| --- | --- | --- | --- |
| Division | 54.627 | 21 | .000 |
| Religion | 55.499 | 9 | .000 |
| Mother's Occupation | 58.544 | 3 | .000 |
| Mother's BMI | 21.187 | 6 | .002 |

From Chi-square test statistics, we have found that the chi-square test statistics are 54.627, 55.499, 58.544, 21.187, and the P-value is .000 ( $<0.05$ ). Hence, we can reject the null hypothesis and say there is an association between the Duration of Breastfeeding and division, religion, mother's occupation, and mother's BMI.

Pearson correlation is a statistical measure that shows the strength and direction of the linear relationship between two continuous variables [39]. Here, we use Pearson Correlation to find how our independent variables (Mother’s age, Mother’s BMI, Total children ever born, Mother’s age at first birth) impact our dependent variable (Duration of Breastfeeding).

**Table 04:** The Pearson’s correlation coefficients between the Duration of Breastfeeding and selected continuous variables.

| Variable | Duration of Breastfeeding Pearson's correlation coefficients | p-value |
| --- | --- | --- |
| Mother's age | 0.113650 | 0.000 |
| Mother's BMI | 0.018620 | 0.193 |
| children ever born | 0.000706 | 0.960 |
| Mothers' age first birth | -0.035651 | 0.012 |

We found a statistically significant positive correlation between the Duration of Breastfeeding and the mother's age ( $r=0.113650$ , $p=0.000$ ), meaning that older mothers tend to breastfeed for more extended periods than younger mothers. There is a statistically significant negative correlation between the Duration of Breastfeeding and the mother's age at first birth ( $r=0.035651$ , $p=0.012$ ), suggesting that mothers who give birth at a younger generation tend to breastfeed for shorter periods than those who have their first child at an older age.

Multinomial logistic regression is a statistical technique used to model the relationship between a categorical dependent variable with three or more categories and one or more independent variables, which can be continuous or categorical [40]. Here, we use the multinomial logistic regression to find the strength and direction of the relationship between the independent and dependent variables (Duration of Breastfeeding). Here, we use the first category of the dependent variable (0-10 months) as a reference category, and for every independent variable, we use the last category as a reference category.

**Table 05:**
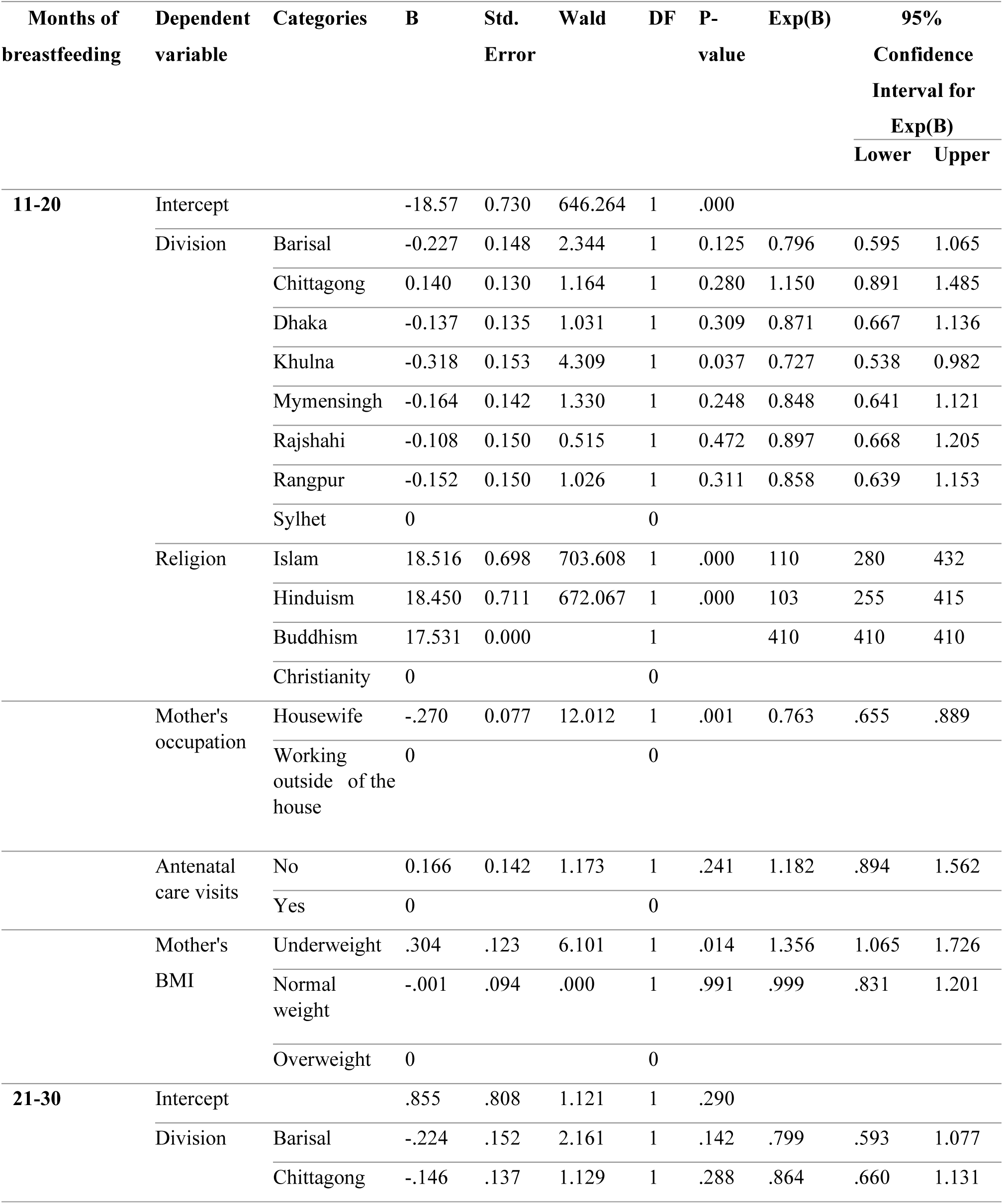
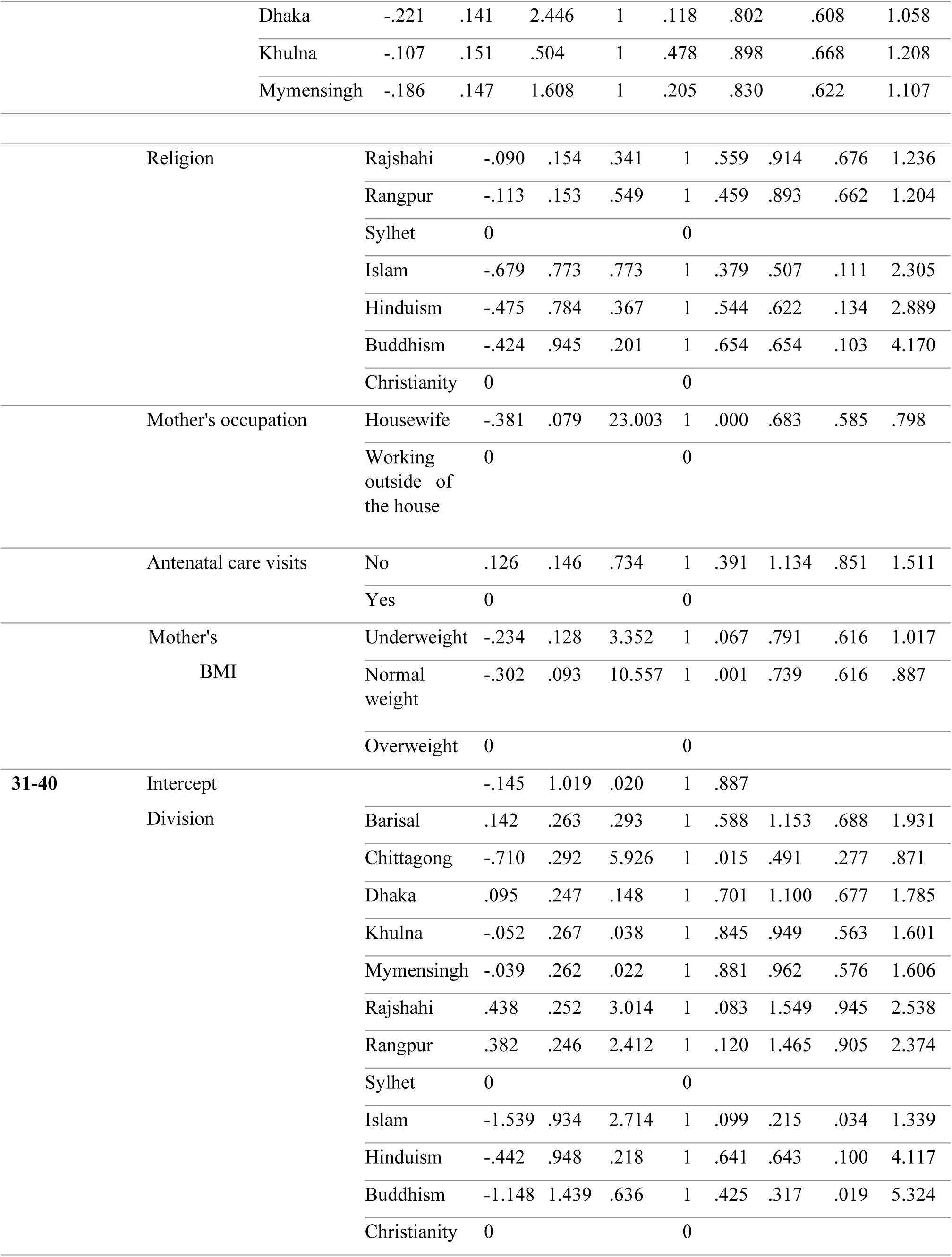
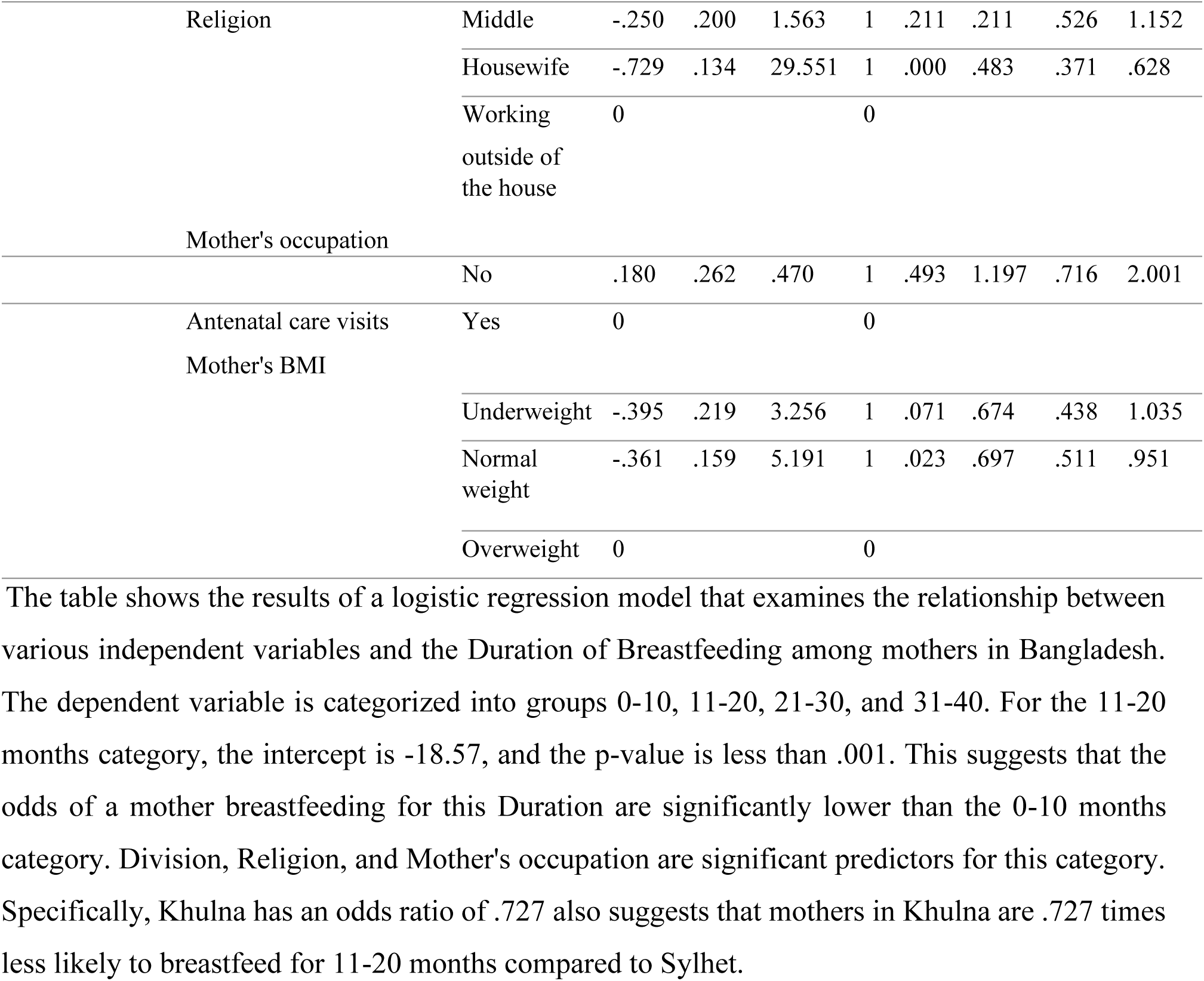
Multinomial logistic regression Parameter Estimates.

Regarding religion, all categories have p-values less than .001, which suggests a significant impact on the Duration of Breastfeeding. Here, the odds ratio of 110 indicates that Muslim mothers are 110 times more likely to breastfeed for 11-20 months than Christian mothers.

The odds ratio of 103 suggests that Hindu mothers are 103 times more likely to breastfeed for 1120 months than Christian mothers.

However, Housewives have an odds ratio of .763, meaning they are less likely to breastfeed for 11-20 months compared to mothers working outside the home, and the p-value is .001. Antenatal care visits and Mother’s BMI do not significantly affect the Duration of Breastfeeding for this category. For the 21-30 months category, the intercept is 0.855, and the p-value is .290, indicating no significant difference in the odds of Breastfeeding compared to the 010 months category. None of the Division categories significantly affect the Duration of Breastfeeding for this category. Regarding religion, all classes have an odds ratio of less than 1 compared to the reference category, indicating that mothers in these categories are less likely to breastfeed for 21-30 months. However, none of the coefficients are significant at the .05 level. Homemakers have an odds ratio of .683, meaning that they are .683 times less likely to breastfeed for this Duration compared to mothers working outside the home, and the p-value is .001, indicating a significant difference.

For BMI, we have found that normal-weighted mothers have an odds ratio of .697, which indicates that normal-weighted mothers are .697 times less likely to breastfeed than over-weighted mothers for 21-30 months. The p-value is .001 for this category, indicating a significant difference. For the 31-40 months category, the intercept is −0.145, and the p-value is .887, indicating no significant difference in the odds of Breastfeeding compared to the 0-10 months category. Chittagong has an odds ratio of .491, indicating that mothers in this region are .491 times less likely to breastfeed for 31-40 months compared to mothers in Sylhet, and the p-value is .015. None of the Religion and Mother’s BMI categories significantly affect the Duration of Breastfeeding for this category. This research found that housewife mothers have an odds ratio of .483, indicating that housewife mothers are .483 times less likely to breastfeed for 31-40 months than mothers working outside. The P-value is .000, which suggests a significant difference.

This research also found that normal-weighted mothers have an odds ratio of .697, indicating that normal-weighted mothers are .697 times less likely to breastfeed for 31-40 months compared to over-weighted mothers. It is worth noting that this category’s Division and Religion variables show different effects compared to the previous two types.

## Result and Discussions

The analysis included 4,881 mothers with at least one child under the age of two years and who had initiated Breastfeeding. The Duration of exclusive Breastfeeding was defined as the length of time between the birth of the child and the introduction of any other liquid or food. Table 01 summarizes the findings of the study. The table shows the mean Duration of Exclusive Breastfeeding (in months), the standard deviation, and the statistical significance of any differences between groups based on various demographic, health-related, socioeconomic, and anthropometric variables.

The study’s results indicate that mothers who received antenatal care had a significantly shorter duration of exclusive breastfeeding than those who did not. This finding is inconsistent with previous studies that have reported a positive association between antenatal care and breastfeeding duration [41]. The authors suggest that this inconsistency may be because mothers who receive antenatal care are more likely to be advised to initiate Breastfeeding early, which may lead to a shorter duration of exclusive Breastfeeding. The study found no significant differences in the Duration of exclusive Breastfeeding between male and female children or between those born via caesarean section versus vaginal delivery. This finding is consistent with previous research that has reported no significant association between the mode of delivery and breastfeeding duration. Mothers who gave birth in a hospital or clinic had a significantly shorter period of exclusive breastfeeding than those who gave birth at home. This finding is consistent with previous studies that have reported a negative association between institutional delivery and breastfeeding duration.

Mothers’ education level was not significantly associated with the Duration of Exclusive Breastfeeding. This finding is inconsistent with previous studies that have reported a positive association between maternal education and breastfeeding duration [42].

Mothers’ occupations had a significant association with the Duration of Exclusive Breastfeeding, with working mothers having a significantly longer duration of exclusive Breastfeeding than homemakers. This finding is consistent with previous studies that have reported a positive association between maternal employment and breastfeeding duration [41].

In addition, the study found that mothers’ education level did not significantly impact the Duration of Exclusive Breastfeeding, with no statistically significant difference observed between the different educational groups. The same was observed for fathers’ education level. However, the mother’s occupation impacted the Duration of exclusive Breastfeeding significantly. Mothers who worked outside the house had a substantially longer duration of exclusive Breastfeeding than housewives. This finding suggests that work policies and support for breastfeeding mothers in the workplace may be necessary for improving breastfeeding rates and infant health.

The study also examined the relationship between the Duration of Exclusive Breastfeeding and the place of residence and wealth index. No significant difference was found in the Duration of exclusive Breastfeeding between mothers living in urban versus rural areas. However, a significant difference was observed in the Duration of exclusive Breastfeeding based on the wealth index, with poor mothers having a longer duration of exclusive Breastfeeding than middle and rich mothers. This finding may be due to poor mothers having less access to formula and other alternatives to Breastfeeding.

Finally, the study found that mothers with a higher BMI (Body Mass Index) had a significantly longer duration of exclusive breastfeeding than those underweight or normal. This finding may be related to the higher nutritional needs of mothers with a higher BMI, as they may be more motivated to breastfeed to ensure adequate nutrition for their infants. Overall, this study provides valuable insights into the factors influencing the Duration of Exclusive Breastfeeding in Bangladesh. The findings suggest that targeted interventions aimed at improving breastfeeding rates and infant health should consider sociodemographic factors such as mothers’ occupation, wealth index, and BMI, in addition to health-related factors such as antenatal care and place of delivery.

It is worth noting that this study has some limitations. Firstly, it is a cross-sectional study, which limits the ability to establish causality between the variables. Additionally, the study relies on self-reported data, possibly subject to recall bias. Future research in this area could benefit from longitudinal study designs and objective measures of breastfeeding duration.

In summary, this study provides important insights into the factors that impact Bangladesh’s Duration of Exclusive Breastfeeding. The findings have important implications for public health policies and interventions to improve breastfeeding rates and infant health in Bangladesh and other low-income countries.

## Conclusion and Future Work

In conclusion, this study aimed to investigate factors influencing breastfeeding duration in Bangladesh. The study collected data from a sample of mothers in Bangladesh and analyzed the Duration of Exclusive Breastfeeding by various demographic, health-related, socioeconomic, and anthropometric variables. The study found that mothers who received antenatal care had a shorter duration of exclusive breastfeeding than those who did not. It is possible that mothers who received antenatal care were advised to supplement their breast milk with formula or other foods, resulting in shorter exclusive breastfeeding durations. The study found no significant difference in the Duration of exclusive Breastfeeding between male and female children or between those born via cesarean section versus vaginal delivery.

In summary, the study found that exclusive Breastfeeding for six months is crucial for ensuring the health of both the mother and child. The study also found that various demographic, health-related, socioeconomic, and anthropometric variables influence the Duration of exclusive breastfeeding. The study’s findings suggest that policymakers and healthcare providers should focus on promoting exclusive Breastfeeding and providing support to mothers who face barriers to Breastfeeding.

Additionally, future research should explore the reasons behind the observed associations between exclusive Breastfeeding and various demographic, health-related, socioeconomic, and anthropometric variables.

## Availability of data and materials

On an adequate request, the corresponding author will make the datasets used and analysed throughout this investigation accessible to the interested party.

## Ethical Approval and Consent to Participate

The authors declare that this manuscript is their original study and is not submitted or under consideration for publication anywhere else.

## Acknowledgment

All authors thank Noakhali Science and Technology University, Bangladesh, for providing various kinds of research support during the preparation of this manuscript.

## Reference

1. Gartner, L.M., et al., Breastfeeding and using human milk. Pediatrics, 2005. 115(2): p. 496–506.

2. Lönnerdal, B., Breast milk: a truly functional food. Nutrition, 2000. 16(7-8): p. 509–11.

3. Gill, S.L., et al., Predicting breastfeeding attrition: adapting the breastfeeding attrition prediction tool. J Perinat Neonatal Nurs, 2007. 21(3): p. 216–24.

4. Li, W., et al., Evaluation of breast milk’s antioxidant capacity and aroma quality. Nutrition, 2009. 25(1): p. 105–14.

5. Gouveri, E., et al., Breastfeeding and diabetes. Curr Diabetes Rev, 2011. 7(2): p. 135–42.

6. James, D.C. and R. Lessen, Position of the American Dietetic Association: promoting and supporting Breastfeeding. J Am Diet Assoc, 2009. 109(11): p. 1926–42.

7. Oddy, W.H., et al., Breastfeeding and cognitive development in childhood: a prospective birth cohort study. Paediatr Perinat Epidemiol, 2003. 17(1): p. 81–90.

8. Hoefer, C. and M.C. Hardy, LATER DEVELOPMENT OF BREAST FED AND ARTIFICIALLY FED INFANTS: COMPARISON OF PHYSICAL AND MENTAL GROWTH. Journal of the American Medical Association, 1929. 92(8): p. 615–619.

9. Bernard, J.Y., et al., Breastfeeding duration and cognitive development at 2 and 3 years of age in the EDEN mother-child cohort. J Pediatr, 2013. 163(1): p. 36–42.e1.

10. Jones, G., et al., How many child deaths can we prevent this year? Lancet, 2003. 362(9377): p. 65–71.

11. Peyre, H., et al., Predicting changes in language skills between 2 and 3 years in the EDENmother–child cohort. PeerJ, 2014. 2: p. e335.

12. Hauck, F.R., et al., Breastfeeding and reduced risk of sudden infant death syndrome: a meta-analysis. Pediatrics, 2011. 128(1): p. 103–10.

13. Kramer, M.S. and R. Kakuma, Optimal Duration of exclusive Breastfeeding. Cochrane Database Syst Rev, 2012. 2012(8): p. Cd003517.

14. Office of the Surgeon, G., et al., Publications and Reports of the Surgeon General, in The Surgeon General’s Call to Action to Support Breastfeeding. 2011, Office of the Surgeon General (US): Rockville (MD).

15. WHO, *Exclusive Breastfeed*.

16. Breastfeeding, S.O., et al., Breastfeeding and the use of human milk. 2012. 129(3): p. e827–e841.

17. NIPORT, National Institute of population research and training (NIPORT), *Mitra and associates, and ICF international. Bangladesh demographic and health survey 2014*. 2013, NIPORT, Mitra and Associates, and ICF International Dhaka, Bangladesh, and ….

18. Haider, R., Kabir, I., Hamadani, J. D., Habibullah, M., & Huttly, S. R., Effect of Breastfeeding on infant and child mortality due to infectious diseases in less developed countries: a pooled analysis. WHO Collaborative Study Team on the Role of Breastfeeding on the Prevention of Infant Mortality. The Lancet, 355(9202), 451–455. 1999.

19. Bernardo, H., V. Cesar, and W.H. Organization, *Long-term effects of Breastfeeding: a systematic review*. 2013.

20. Victora, C.G., et al., Breastfeeding in the 21st century: epidemiology, mechanisms, and lifelong effect. 2016. 387(10017): p. 475–490.

21. Stuebe, A.J.R.I.o. and gynecology, The risks of not breastfeeding for mothers and infants. 2009. 2(4): p. 222.

22. Binns, C., et al., Guidelines for complementary feeding of infants in the Asia Pacific region: APACPH Public Health Nutrition Group. 2020. 32(4): p. 179–187.

23. Saha, K.K., Frongillo, E. A., Alam, D. S., Arifeen, S. E., & Persson, L. A., Racial/ethnic and education-related disparities in the Duration of Breastfeeding among women in Bangladesh. Journal of Human Lactation, 2008. 24(4), 377–386.

24. Bhutta, Z.A., et al., What works? Interventions for maternal and child undernutrition and survival. 2008. 371(9610): p. 417–440.

25. Organization, W.H., *Maternal, newborn, child, and adolescent health: Bangladesh*. 2020.

26. WHO, U., Progress on Breastfeeding in Bangladesh undermined by aggressive formula milk marketing. 23 February 2022.

27. Giashuddin, M.S. and M. Kabir, Duration of Breastfeeding in Bangladesh. Indian J Med Res, 2004. 119(6): p. 267–72.

28. Doan, T.T.D., et al., Improving Breastfeeding by empowering mothers in Vietnam: A randomized controlled trial of a mobile app. 2020. 17(15): p. 5552.

29. Rollins, N.C., et al., Breastfeeding 2: why invest, and what it will take to improve breastfeeding practices. 2016. 387(10017): p. 491–504.

30. Edhborg, M., et al., Impact of postnatal maternal depressive symptoms and infant’s sex on mother-infant interaction among Bangladeshi women. 2013. 5(02): p. 237.

31. Anik, A.I., et al., Double burden of malnutrition at household level: A comparative study among Bangladesh, Nepal, Pakistan, and Myanmar. 2019. 14(8): p. e0221274.

32. Khan, J., et al., Timing of breastfeeding initiation and exclusivity of Breastfeeding during the first month of life: effects on neonatal mortality and morbidity—a systematic review and meta-analysis. 2015. 19: p. 468–479.

33. Ahmed, A.S., Khanam, P. A., & Parveen, S., Practices of traditional postpartum confinement among the urban poor of Dhaka, Bangladesh: a qualitative study. BMC pregnancy and childbirth, 2010. 10(1), 79.

34. Mamun, A.A., & Lawlor, D. A, Infant feeding and maternal health behaviors influence the incidence of diarrhea and nutritional status of young children in Bangladesh. Public health nutrition, 2012. 15(9), 1691–1700.

35. QK, A., Socio-economics of Bangladesh through the decades. 2018, Dhaka: Pathak Shamabesh.

36. Allen, C., J. Fleuret, and J. Ahmed. Data quality in demographic and health surveys that used long and short questionnaires. 2020. ICF.

37. ICF, T.D.P. and M. Rockville, U.S.A., Bangladesh Demographic and Health Survey 2017–18. 2019.

38. Tallarida, R.J., et al., Chi-square test. 1987: p. 140–142.

39. Cohen, I., et al., Pearson correlation coefficient. 2009: p. 1–4.

40. Kwak, C. and A.J.N.R. Clayton-Matthews, Multinomial logistic regression. 2002. 51(6): p. 404–410.

41. Khanal, V., et al., Factors associated with early initiation of Breastfeeding in Western Nepal. 2015. 12(8): p. 9562–9574.

42. Alemayehu, T., J. Haidar, and D.J.E.J.o.H.D. Habte, Determinants of exclusive breastfeeding practices in Ethiopia. 2009. 23(1).

